# Benchmarking Speech Recognition Models for Medical Consultations in Latin American Spanish: A Comparative Evaluation with Fine-Tuning

**DOI:** 10.64898/2026.07.14.26358062

**Authors:** Rodrigo M. Carrillo-Larco, Ariana Carbajal Serrano, Paulo S Condori Pinedo

## Abstract

**BACKGROUND:** Artificial intelligence (AI) medical scribes rely on speech-to-text (STT) models for transcription. Evaluations of STT models in non-English settings remain scarce. We benchmarked ten STT models on medical consultations from Latin American (LatAm) Spanish and assessed whether fine-tuning improves transcription accuracy.

**METHODS:** Ten videos depicting medical consultations. Human transcriptions were the ground truth. Five closed-source models were evaluated: gpt-4o-transcribe, gpt-4o-mini-transcribe, gemini-2.5-pro, Eleven Labs, and Assembly AI; and so were five open-source models: Whisper Large, Whisper Large v3, Whisper Large v3 Turbo, Voxtral Mini 3B, and Canary 1B v2. Whisper Large v3, an open-source model, was fine-tuned. One video was withheld from training. Performance assessed using Word Error Rate (WER), Character Error Rate (CER), BLEU Score, ROUGE-L, BERT Score, and Semantic Similarity on the one withheld video.

**RESULTS:** None of the fine-tuning iterations outperformed the vanilla Whisper Large v3. When evaluated on the held-out test video, the fine-tuned open-source Whisper Large v3 did not outperform any of the top closed-source models; conversely, relative to other open-source models, the fine-tuned Whisper Large v3 had superior performance across all metrics, such as in BLEU score (63% vs. 58% for the second-best model), BERT Score (89% vs. 86%), and semantic similarity (89% vs. 83%).

**CONCLUSIONS:** Whisper Large v3 and its fine-tuned variant are the best open-source STT models for transcribing medical conversations in LatAm Spanish. These findings provide evidence for developing AI medical scribes tailored to Spanish-speaking LatAm.

## INTRODUCTION

Artificial intelligence (AI) applications in healthcare have evolved from prediction models based on structured tabular data and medical imaging to large language models (LLMs) capable of text summarization and clinical question-answering.^1,2^ Alongside LLMs, speech-to-text (STT) models, also known as automatic speech recognition (ASR) models, have emerged as tools for automatizing clinical documentation.^3,4^ These systems, commonly referred to as AI medical scribes, can record a clinical encounter, produce a verbatim transcription, and generate a clinical note. By automating this process, AI medical scribes hold the promise of reducing the time physicians spend on documentation and of improving the overall quality of clinical interactions.^5–12^

The potential of AI medical scribes is reflected in a growing body of academic literature as well as in the rapid expansion of commercial offerings in this space. Recent clinical trials have suggested that medical scribes can reduce physician stress and documentation time without compromising quality.^5,6^ The proliferation of both research activity and commercial products underscores the need for rigorous, context-specific evaluations of AI scribe performance. Central to any such evaluation is the STT component, which determines the accuracy of the transcript from which the clinical note is derived. Errors introduced at the transcription stage propagate through the documentation pipeline and can compromise the safety and reliability of the resulting clinical note.

Systematic evaluation of STT models in non-English settings remains limited.^13,14^ Whether current STT systems can reliably transcribe medical conversations conducted in Spanish is largely unknown. This knowledge gap is consequential for Latin American (LatAm) Spanish, which is characterized by phonological and lexical variation across the region. Beyond dialectal diversity, LatAm clinical speech poses additional challenges: the epidemiological landscape of the region covers a mixture of chronic, infectious, and tropical diseases,^15,16^ which generates specialized terminology. Other factors that may affect STT performance include variability in patient education level, the use of colloquial expressions to describe symptoms and illnesses, health beliefs that shape the content of clinical discourse, and suboptimal recording conditions in some healthcare settings. Characterizing STT model performance across this heterogeneous context is a prerequisite for developing reliable AI medical scribes for LatAm.

We aimed to benchmark several STT models on medical consultation recordings in LatAm Spanish and to determine whether fine-tuning one open-source STT model on a domain-specific dataset could yield superior transcription accuracy. We evaluated performance across several ASR metrics relative to human-generated transcriptions. We assessed whether fine-tuning reduced performance variability, important for reliable deployment in clinical settings.

## METHODS

### Data Sources

We assembled a dataset of 10 videos depicting medical consultations conducted in Spanish from five LatAm countries and one from the United States, retrieved from YouTube (Supplementary Table 1). All videos were freely accessible without login credentials or subscriptions. Videos were downloaded as audio-only files. A human transcriber with experience in qualitative research transcription produced verbatim transcriptions of each recording. A second reviewer independently verified all transcriptions for accuracy. The human transcriptions were the gold-standard reference against which STT models were evaluated.

### Montreal Forced Aligner

Fine-tuning an STT model requires short audio segments, typically 10 to 15 seconds in duration, paired with their corresponding reference transcriptions. To create this training corpus, we used the Montreal Forced Aligner (MFA) to segment each full-length video into ∼10-second audio chunks and to align each segment with the appropriate portion of the human transcription. Despite exhaustive attempts with varying hyperparameters, one video (v00003) could not be processed by the MFA algorithm and was excluded from fine-tuning. This video (v00003) was retained as a fully independent external validation set, having played no role whatsoever in model training.

Processing the nine eligible videos with the MFA yielded 594 audio segments (Supplementary Table 2). The mean segment duration was 10.7 seconds (standard deviation (SD): 2.52 seconds). Segments with durations exceeding 10 seconds generally arose from extended periods of silence, such as those occurring during physical examination maneuvers, including when a physician paused to auscultate the patient’s heart during which no speech was present. The aggregate duration of the 594-segment dataset was 6,356.51 seconds (105.94 minutes). A human reviewer confirmed the accuracy of the MFA-generated audio-to-text pairings.

### Data Augmentation

For each original chunk (n=594), four augmented variants were generated, yielding a final dataset of 2,970 audio samples: 594 original recordings plus 2,376 (594 x 4) augmented samples. All augmented samples preserved their original transcription.

Four augmentation regimes were implemented using the *Audiomentations* library in Python. The first regime (“noisy clean”) introduced low-amplitude Gaussian noise, mild pitch perturbations, and small gain variations. The second regime (“multi-speaker”) overlayed an unrelated speech segment from a different recording at a randomly selected signal-to-noise ratio between 8 and 18 dB, followed by additional low-level microphone hiss. The third regime (“acoustic environment”) added colored noise and synthetic reverberation. Pink-to-brown spectral noise was injected at stochastic signal-to-noise ratios, after which the signal was convolved with a synthesized exponentially decaying impulse response. The fourth regime (“broad combination”) applied a heavier stack of perturbations, including Gaussian noise, pitch shifting, temporal stretching, gain variation, temporal shifting, colored noise, and 30% probability of synthetic reverberation.

### Analysis Plan

#### Benchmarking STT models

We benchmarked five closed-source models: gpt-4o-transcribe; gpt-4o-mini-transcribe; gemini-2.5-pro; Eleven Labs (scribe_v2); and Assembly AI. We benchmarked five open-source models: Whisper Large; Whisper Large v3; Whisper Large v3 Turbo; Voxtral Mini 3B; and Canary 1B v2.

Open-source models encompass state-of-the-art versions developed by major AI companies, including OpenAI and Google. Open-source model selection followed two criteria. First, all selected models are available through Hugging Face under the Apache 2.0 license, permitting commercial and non-commercial applications. Second, these five models occupied the top five positions on the Hugging Face Speech Recognition Multilingual Leaderboard at the time of the analysis (January 2026), ranked by the lowest WER. While all five models can process Spanish-language audio, none has been specifically optimized for medical speech from LatAm.

#### Evaluation Metrics

Six metrics were used to assess transcription quality against the human reference. Word Error Rate (WER) measures the proportion of words in the reference transcription that were incorrectly predicted by the model and is the standard primary metric in automatic speech recognition. Character Error Rate (CER) applies the same edit-distance logic at the character level, making it more sensitive to partial word errors and morphological variation. BLEU score (Bilingual Evaluation Understudy) quantifies n-gram overlap between the model output and the reference transcription, rewarding sequences of consecutive words that match the gold standard. ROUGE-L (Recall-Oriented Understudy for Gisting Evaluation – Longest Common Subsequence) captures the longest common subsequence between the transcription and reference, reflecting fluency and order preservation even when exact n-gram matches are absent. BERT Score leverages contextual embeddings from a pretrained BERT model to compute token-level similarity between transcription and reference, capturing semantic equivalence. Semantic Similarity was computed using the *sentence-transformers/paraphrase-multilingual-MiniLM-L12-v2 model*, the top-ranked multilingual sentence transformer at the time of analysis, which encodes both the transcription and reference as dense vectors and measures their cosine similarity, providing a holistic, language-agnostic estimate of meaning preservation. For all metrics except WER and CER, values closer to 1.0 indicate superior performance.

Although WER and CER are the standard ASR metrics, their word- and character-level matching can misrepresent the clinical utility of a transcript; for instance, paraphrase, synonymy, and format variation are penalized as if they were errors (e.g., “*paciente refiere*” vs. “*paciente reporta*”, both rendered in English as “the patient reports”, would be treated by as substitutions despite being equivalent). The other metrics address these limitations. BLEU rewards correct, fluent n-grams. ROUGE-L captures content recall via the longest common subsequence. BERT Score assesses whether tokens are semantically equivalent to the reference even if surface forms differ. Semantic Similarity quantifies meaning preservation.

#### Fine-Tuning

Whisper Large v3, an open-source model, was selected for fine-tuning based on its status as the most capable and widely adopted recent iteration of the Whisper model family. The training procedure followed a nine-fold leave-one-out (LOO) cross-validation design across nine eligible videos, with a tenth video (v00003) fully withheld for external validation. In each fold, the model was trained on audio segments from eight videos and evaluated on the complete, full-length recording of the single held-out video. Within each training fold, the available data were further partitioned into an 80% training subset and a 20% validation subset to monitor for overfitting and to implement early stopping with patience of three evaluation steps monitored via evaluation loss. Fixed training hyperparameters included a batch size of 2, a maximum of 50 epochs, 20 warm-up steps, evaluation and checkpoint-saving intervals of every 10 steps, and a gradient accumulation factor of 2. Three learning rates were explored to optimize performance: 1e-5, 5e-6, and 2e-5. Beam sizes of 3, 5 (default), and 7 were tested during inference to evaluate the trade-off between decoding speed and accuracy.

LOO validation results were summarized as the mean and SD across the nine folds. The best-performing hyperparameter configuration (learning rate and beam size) was selected based on summary metric performance and applied to produce the fine-tuned, production-ready model.

### Statistical Analysis

The statistical analysis was descriptive and aimed at characterizing the performance of both the benchmarked models and the fine-tuned model. A hypothesis was formulated that the fine-tuned model would demonstrate superior performance compared to the benchmarked models when applied to a single video (v00003), which was not utilized during the fine-tuning process. Although this comparison is limited to a single video, it serves as a fair evaluation because the fine-tuned model was benchmarked against a completely new and previously unseen video. If all videos had been utilized for benchmarking, the fine-tuned model would have shown unrealistically better performance because it was being evaluated with nine videos that were used in its fine-tuning.

First, to establish a baseline, we applied the ten STT models (closed-source and open-source) to the ten videos and computed the performance metrics. These metrics were summarized across the ten videos as means and standard deviations. Second, we summarized the performance metrics across the nine folds in the LOO validation during the fine-tuning phase. We presented the mean and standard deviation for each metric and hyperparameter variation (learning rate and beam number) across the nine folds. Third, we applied the ten vanilla STT models as well as the fine-tuned model to the withheld video (video 00003 was not utilized during fine-tuning). We computed the validation metrics and ranked the model’s performance across the metrics. Importantly, because this last evaluation was conducted using a single video (v00003), model rankings and performance metrics may vary when models are evaluated on larger cohorts, diverse regional accents, varied acoustic environments, or different clinical specialties.

### Contributions

From a technical perspective, our core contribution lies not in architectural novelty because our pipeline follows a standard workflow of data collection, fine-tuning, and multi-model benchmarking, but in its domain application. To the best of our knowledge, this work is the first systematic evaluation of STT models using LatAm Spanish. Furthermore, we contribute a novel resource by sourcing audio from YouTube, similar to prior work.^14^ This uncurated, real-world dataset introduces acoustic noise and variability, providing a rigorous and realistic benchmark for STT performance.

### Ethics

This study used publicly available videos that do not require privileged access. No human subjects were directly involved, and no personally identifiable or clinically sensitive information was collected or disclosed. The research was deemed as minimal-risk, non-human-subjects research.

### Role of the Funding Source

No funding was available for this project. The views expressed herein represent those of the authors alone and do not necessarily reflect the positions of their respective affiliated institutions.

### AI Use Disclosure

Claude Sonnet 4.6 (Anthropic) and Gemini 3.5 Flash were used to assist with manuscript editing and abstract preparation. All AI-assisted revisions were reviewed and approved by the authors, who accept full responsibility for the accuracy and integrity of the manuscript’s content.

## RESULTS

### Vanilla STT Models on 10 Full-Length Videos

In the evaluation of the ten full-length videos, closed-source models showed superior performance compared to open-source models (Table 1). Gemini-2.5-pro (closed-source) emerged as the leading model, surpassing most metrics, including WER at 14.0%, CER at 9.0%, and semantic similarity at 95.8%. Within the open-source models, Whisper Large v3 consistently outperformed its counterparts, achieving WER of 18.6%, CER of 11.8%, and semantic similarity of 93.4%. Thus, Whisper Large v3 underwent fine-tuning.

**Table 1.** Performance of the Vanilla Models Applied to the Ten Full-Length Videos.

| Model | WER |  | CER |  | BLEU Score |  | ROUGE-L |  | BERT Score |  | Semantic Similarity |  |
| --- | --- | --- | --- | --- | --- | --- | --- | --- | --- | --- | --- | --- |
|  | Mean | STD | Mean | STD | Mean | STD | Mean | STD | Mean | STD | Mean | STD |
| **Closed-source models** |  |  |  |  |  |  |  |  |  |  |  |  |
| **gpt-4o-transcribe** | 0.297 (4) | 0.168 (4) | 0.224 (4) | 0.118 (4) | 0.603 (4) | 0.198 (4) | 0.807 (4) | 0.125 (4) | 0.925 (1) | 0.020 (1) | 0.956 (2) | 0.027 (2) |
| **gpt-4o-mini-transcribe** | 0.420 (5) | 0.303 (5) | 0.340 (5) | 0.246 (5) | 0.542 (5) | 0.245 (5) | 0.717 (5) | 0.213 (5) | 0.918 (4) | 0.036 (4) | 0.953 (3) | 0.040 (4) |
| **gemini-2.5-pro** | 0.140 (1) | 0.058 (1) | 0.090 (1) | 0.042 (1) | 0.780 (1) | 0.074 (2) | 0.913 (1) | 0.037 (1) | 0.922 (2) | 0.029 (3) | 0.958 (1) | 0.031 (3) |
| **Eleven Labs (scribe_v2)** | 0.170 (3) | 0.072 (3) | 0.118 (3) | 0.056 (3) | 0.742 (3) | 0.082 (3) | 0.896 (3) | 0.045 (2) | 0.883 (3) | 0.037 (5) | 0.933 (4) | 0.063 (5) |
| **Assembly AI** | 0.156 (2) | 0.067 (2) | 0.105 (2) | 0.050 (2) | 0.769 (2) | 0.073 (1) | 0.897 (2) | 0.047 (3) | 0.922 (2) | 0.021 (2) | 0.958 (1) | 0.027 (1) |
| **Open-source models** |  |  |  |  |  |  |  |  |  |  |  |  |
| **Whisper Large** | 0.578 (3) | 0.480 (5) | 0.402 (3) | 0.326 (4) | 0.456 (3) | 0.335 (5) | 0.560 (3) | 0.343 (5) | 0.893 (2) | 0.047 (3) | 0.940 (2) | 0.053 (1) |
| **Whisper Large-v3-Turbo** | 0.196 (2) | 0.091 (3) | 0.122 (2) | 0.064 (3) | 0.708 (2) | 0.106 (4) | 0.866 (2) | 0.065 (4) | 0.888 (3) | 0.044 (2) | 0.913 (3) | 0.083 (3) |
| **Whisper Large v3** | 0.186 (1) | 0.079 (1) | 0.118 (1) | 0.057 (2) | 0.722 (1) | 0.097 (3) | 0.874 (1) | 0.057 (2) | 0.910 (1) | 0.033 (1) | 0.934 (1) | 0.060 (2) |
| **Voxtral Mini 3B** | 1.546 (5) | 0.408 (4) | 1.172 (5) | 0.406 (5) | 0.029 (4) | 0.019 (2) | 0.118 (4) | 0.058 (3) | 0.662 (4) | 0.051 (4) | 0.777 (4) | 0.143 (5) |
| **Canary 1B v2** | 0.956 (4) | 0.027 (2) | 0.864 (4) | 0.055 (1) | 0.006 (5) | 0.007 (1) | 0.081 (5) | 0.045 (1) | 0.636 (5) | 0.058 (5) | 0.697 (5) | 0.119 (4) |

CER = Character Error Rate; FT = fine-tuned model; SD = standard deviation; WER = Word Error Rate. Values represent means and standard deviations across the 10 videos. For WER and CER, lower values indicate better performance; for BLEU Score, ROUGE-L, BERT Score, and Semantic Similarity, values closer to 1.0 indicate better performance. The numerical value enclosed in parentheses (1-5) signifies the ranking, with 1 denoting the highest and 5 the lowest within each group of models (closed-source and open-source).

### Fine-Tuned Model (Whisper Large v3, an open-source model)

None of the fine-tuning iterations outperformed the vanilla Whisper Large v3 (Table 2). Furthermore, fine-tuning with data augmentation resulted in worse performance compared to fine-tuning with*out* data augmentation. Consequently, further experiments were conducted without data augmentation.

**Table 2.**
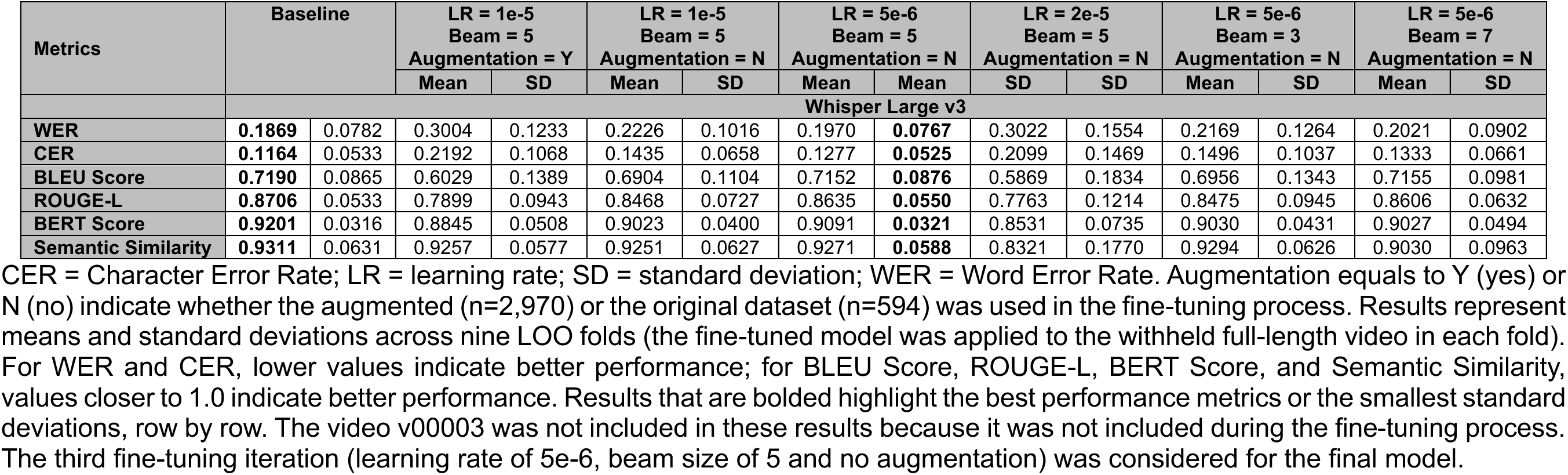
Validation Metrics During Fine-Tuning, Internal Leave-One-Out (LOO) Validation.

The fine-tuning configuration that used a learning rate of 5e-6, a beam size of 5, and no data augmentation, achieved the closest metrics in comparison to the vanilla model. For instance, the WER of the fine-tuned model was 19.7% (18.6% vanilla), the CER was 12.7% (11.6% vanilla), and the semantic similarity was 92.7% (93.1% vanilla).

### Benchmarking the Fine-Tuned open-source STT Model in the Withheld Video

When evaluated on the held-out test video (v00003), the fine-tuned open-source Whisper Large v3 model did not outperform any of the top closed-source models (Table 3, upper panel). Notably, Whisper Large v3 ranked fourth out of six models across five of the six metrics, showing superior performance relative to two closed-source models.

**Table 3.** Performance of the models applied to the withheld video only.

| Model | WER | CER | BLEU Score | ROUGE-L | BERT Score | Semantic Similarity |
| --- | --- | --- | --- | --- | --- | --- |
| **Closed-source models** |  |  |  |  |  |  |
| **gpt-4o-transcribe** | 0.6756 (6) | 0.6538 (6) | 0.1558 (5) | 0.4993 (6) | 0.9101 (2) | 0.9543 (3) |
| **gpt-4o-mini-transcribe** | 0.6728 (5) | 0.6465 (5) | 0.1547 (6) | 0.5041 (5) | **0.9121** (1) | 0.9726 (2) |
| **gemini-2.5-pro** | **0.2078** (1) | **0.1465** (1) | **0.6999** (1) | **0.8715** (1) | 0.8608 (5) | 0.8866 (6) |
| **Eleven Labs (scribe_v2)** | 0.2190 (2) | 0.1515 (2) | 0.6764 (2) | 0.8570 (2) | 0.8504 (6) | 0.9175 (4) |
| **Assembly AI** | 0.2334 (3) | 0.1612 (3) | 0.6703 (3) | 0.8379 (3) | 0.9043 (3) | **0.9795** (1) |
| **Whisper Large v3 (Fine-tuned)** | 0.2933 (4) | 0.1967 (4) | 0.6309 (4) | 0.7966 (4) | 0.8913 (4) | 0.8987 (5) |
| **Open-source models** |  |  |  |  |  |  |
| **Whisper Large** | 0.7818 (4) | 0.5751 (4) | 0.1903 (4) | 0.2541 (4) | 0.8639 (2) | 0.8328 (2) |
| **Whisper Large-v3-Turbo** | 0.2996 (3) | 0.2120 (3) | 0.5778 (3) | 0.7912 (3) | 0.8309 (4) | 0.7185 (4) |
| **Whisper Large v3** | 0.2963 (2) | 0.2049 (2) | 0.5805 (2) | 0.7931 (2) | 0.8420 (3) | 0.8202 (3) |
| **Voxtral Mini 3B** | 0.9863 (5) | 0.7021 (5) | 0.0016 (5) | 0.0665 (5) | 0.6046 (5) | 0.6215 (5) |
| **Canary 1B v2** | 0.9995 (6) | 0.9048 (6) | 0.0000 (6) | 0.0019 (6) | 0.5380 (6) | 0.4810 (6) |
| **Whisper Large v3 (Fine-tuned)** | **0.2933** (1) | **0.1967** (1) | **0.6309** (1) | **0.7966** (1) | **0.8913** (1) | **0.8987** (1) |

CER = Character Error Rate; WER = Word Error Rate. Video v00003 was excluded from all stages of model fine-tuning and serves as a fully external validation set. For WER and CER, lower values indicate better performance; for BLEU Score, ROUGE-L, BERT Score, and Semantic Similarity, values closer to 1.0 indicate better performance. Results that are bolded highlight the best performance metrics column-wise. The numerical value enclosed in parentheses (1-6) signifies the ranking, with 1 denoting the highest and 6 the lowest. Importantly, because this evaluation was conducted using a single consultation transcript (v00003), model rankings and performance metrics may vary when evaluated across larger cohorts, diverse regional accents, varied acoustic environments, or different clinical specialties.

Conversely, relative to the other open-source models, the fine-tuned Whisper Large v3 model showed superior performance across all evaluated metrics (Table 3, lower panel). The most pronounced improvements were seen in BLEU score (63% vs. 58% for the second-best model), BERT Score (89% vs. 86%), and semantic similarity (89% vs. 83%).

## DISCUSSION

### Main Findings

We curated a dataset of videos depicting medical consultations in LatAm Spanish and evaluated five closed-source and five open-source STT models alongside a domain-specific fine-tuned open-source model, benchmarking all against human transcriptions as the gold standard. Among the vanilla models, Gemini-2.5-pro (closed-source) and Whisper Large v3 (open-source) achieved the best performance across all ten videos. In the external validation analysis, restricted to a single video withheld from the fine-tuning process, the fine-tuned model ranked first in all metrics among open-source models, yet the fine-tuned open-source model ranked fourth among the closed-source models.

Taken together and despite the inherent limitations of a small dataset sourced from publicly available online recordings, these findings provide the first characterization of STT models in the LatAm Spanish medical setting. These findings suggest that Gemini-2.5-pro is the best closed-source STT model, whereas, amongst open-source models, Whisper Large v3 as well as its fine-tuned version offer a strong baseline for medical applications in LatAm Spanish. Broader and more diverse evaluations will be needed to validate these observations and to develop STT models that can be deployed in real-world clinical scenarios in Spanish-speaking LatAm.

### Strengths and Limitations

This study has many strengths. We rigorously and transparently evaluated STT models against human-derived gold-standard transcriptions. We included both closed-source as well as open-source STT models. The fine-tuning procedure employed a LOO cross-validation design to minimize overfitting, and the model’s generalizability was further examined on an independent external recording that was completely excluded from all stages of training. The dataset incorporated videos from multiple countries and diverse clinical contexts, introducing meaningful variability in regional accents, colloquial expressions, and clinical terminology.

Notwithstanding, limitations must be acknowledged. First, the study dataset was small, comprising 10 videos in total, of which nine were used for fine-tuning. Although the inclusion of geographically diverse recordings is a strength, 10 consultations fall short of the hundreds of hours typically required to train robust STT models for specialized domains. This constraint likely attenuated the performance of the fine-tuned model, which, while superior to other open-source models, did not outperform closed-source models. Expanded datasets incorporating greater variation in regional accents, colloquialisms, and clinical scenarios, would be expected to yield larger performance gains. Work is currently underway to enlarge the dataset through additional recordings and through the generation of synthetic data, wherein LLMs produce scripted clinical dialogues that are subsequently rendered as audio by a text-to-speech model. Second, the small dataset permitted only a single recording for external validation. A single data point does not support meaningful estimation of variability in an independent dataset. Third, the videos utilized, while helpful and constituting the first dataset of this nature, did not always depict the entirety of a medical consultation (e.g., introduction, medical history, physical examination, prescription, and explaining the prescription to the patient). Consequently, our research was unable to transcribe and evaluate the complete intricacies of a complete medical consultation. Fourth, while the closed-source STT models benchmarked in this study represent state-of-the-art general-purpose systems, they are not specifically optimized for clinical contexts, unlike the commercial AI medical scribes currently deployed in healthcare settings. This is a limitation because our findings cannot assess the performance of medical AI scribes when processing LatAm Spanish clinical encounters. Fifth, our evaluation relied primarily on error metrics such as WER; however, clinical utility requires assessing the precise extraction of domain-specific entities, including signs, symptoms, diagnoses, medications, and therapeutic procedures using metrics like precision, recall, and F1 score. Conducting such entity-level evaluations requires extensive manual annotation of clinical transcripts, a process that was beyond the scope of this initial benchmark.

Future endeavors should seek to obtain videos or audios of a complete medical consultation; it is paramount to collect a (much) larger dataset. Regarding technical innovation, future research should explore emerging STT architectures and focus on developing novel modeling strategies rather than relying solely on fine-tuning and benchmark evaluation. Similarly, future research should prioritize rigorous, independent evaluations of commercial AI medical scribes currently being deployed in predominantly LatAm Spanish-speaking clinical settings.

### Implications

Other evaluations in non-English languages have similarly demonstrated strong, though non-optimal, performance metrics. For instance, previous work in Vietnamese^14^ reported a WER of 26.8%, while benchmarking in French^13^ achieved a best WER of 17.21%. Our fine-tuned model achieved a best WER of 29.3%. This higher WER is likely due to dataset volume constraints because these studies leveraged larger datasets: >240 hours in French^13^ and 16 hours of labeled medical speech in Vietnamese^14^.

This study carries meaningful implications for research, clinical practice, and health technology development. On the research side, we contribute with the first curated dataset of medical consultation recordings in LatAm Spanish designed for ASR evaluation (Supplementary Table 1). However, its small size (n=10) underscores the urgent need for data collection efforts across LatAm to produce corpora of sufficient size and diversity to support large-scale STT model development and validation, as it is happening in other regions such as Africa.^17,18^

From a clinical and implementation standpoint, our findings have utility for practitioners and technology companies seeking to deploy AI medical scribes in Spanish-speaking LatAm. The lead of a fine-tuned version of Whisper Large v3 (open-source model) over the other evaluated baselines provides an evidence-based starting point for any development team building a LatAm Spanish medical scribe with open-source STT models. The gains achieved through fine-tuning, even with a small dataset, suggest that domain adaptation is a high-value strategy: investments in curating domain-specific training data can yield improvements in both transcription accuracy and reliability.

Our results also carry broader public health implications. Access to accurate AI-assisted clinical documentation tools could meaningfully address physician burnout in a region where healthcare systems are often under-resourced and primary care physicians bear substantial loads.^19^ If fine-tuned STT models can reliably capture the nuances of LatAm clinical Spanish, they could enable the development of AI scribes that reduce documentation time and allow clinicians to devote more attention to patient care and themselves.^5,6^ This is particularly consequential in settings where support staff are limited, and physicians bear the full burden of clinical documentation.

Finally, from a methodological perspective, our framework offers a reproducible template that investigators can adapt for other underrepresented languages and clinical contexts (benchmarking closed-source and open-source models, fine-tuning the best-performing candidate, and evaluating on a fully withheld external set).

### Interpretation of Main Findings

An intriguing finding is that fine-tuning Whisper Large v3 (open-source model) on the augmented dataset yielded worse performance metrics compared to training on the non-augmented data. This finding appears counterintuitive, as conventional paradigms assume that expanding dataset scale and diversity improves model generalization. We propose the following hypotheses to explain these findings; however, these interpretations are speculative and warrant empirical validation. In the context of state-of-the-art STT foundation models, this degradation highlights a complex interplay between heavy perturbation, domain shifting, and the architectural robustness of the underlying model. First, this performance drop underscores a potential optimization conflict regarding regularization overkill on a pre-trained foundation model. Whisper Large v3^20^ (open-source model) has already been exposed to millions of hours of highly diverse and multi-lingual audio during its training phase. Consequently, it already has intrinsic robustness to acoustic variation. When fine-tuning, forcing it to adapt to aggressive synthetic data, rather than teaching the model novel acoustic concepts, this process likely disrupts high-quality pre-trained weights. Second, by aggressively injecting synthetic artifacts, the model’s attention mechanism was conditioned to decode through artificial distortion. The discrepancy in acoustic distributions meant that the model was optimized for a noisy target that did not align with the characteristics of the actual evaluation recordings, ultimately elevating the WER during inference. Consequently, fine-tuning with non-augmented data delivered better results because the baseline foundation model was spared from large distortions. Rather than forcing the model’s attention layers to adapt to artificial artifacts, training solely on the clean signal allowed the model to preserve its highly optimized pre-trained weights. This likely focused the fine-tuning process on the linguistic and phonetic boundaries of the target domain, allowing the model to adapt smoothly to specialized terminology without inducing catastrophic forgetting or optimization instability. We reiterate that these hypotheses remain speculative, and further empirical evaluation is required to ascertain the underlying mechanisms driving these findings.

A primary constraint on model performance following fine-tuning was dataset volume. We hypothesize that expanding the training data, with or without targeted data augmentation, would yield gains in recognition metrics. Systematically testing this scaling hypothesis would require fine-tuning across incrementally meaningful data regimes (e.g., 100, 500, and 1,000 hours). However, given our current dataset size (∼106 minutes), conducting a meaningful scaling analysis was beyond the scope of this study.

## Conclusions

Whisper Large v3 (open-source model) and its fine-tuned version represent the most accurate open-source STT models identified for transcribing medical conversations in Spanish from LatAm, while Gemini-2.5-pro (closed-source model) was the closed-source model with the best performance. These findings provide an empirical foundation for developing AI medical scribe applications tailored to the linguistic and clinical diversity of Spanish-speaking LatAm. As generative AI technologies continue to expand into the LatAm, rigorous, context-specific evaluations are essential to ensure that these tools meet the performance standards required for safe and effective clinical use across the heterogeneous populations and clinical scenarios of LatAm.

## Data Availability

The data utilized in this study (videos depicting medical consultations in Latin American Spanish) are freely accessible online. Supplementary Table 1 provides direct links to these videos. However, we do not possess the permissions to further disseminate these materials; thus, we are not making the videos or transcriptions open access.

## DECLARATIONS

## ACKNOWLEDGEMENTS

With Claude (Sonnet 4.6) and Gemini 3.5 Flash we enhanced the manuscript and generated ideas for the discussion. The authors composed the initial draft of the manuscript and submitted it to Claude and Gemini for editing and refinement, with the objective of enhancing clarity and impact. Then, the authors verified the content. The authors collectively bear full responsibility for the content of the manuscript. All the models tested herein were executed on the HyPER C3 – Community Cloud HPC Cluster at Emory University, Atlanta, USA.

## FUNDING

No specific funding was available for this project.

## CONFLICTS OF INTEREST

The authors are utilizing these findings and the fine-tuned STT model to develop a commercially viable product for the Latin American market. The fine-tuned model cannot be shared; similarly, the human transcriptions (gold standard or ground truth used in the evaluations) cannot be shared.

## CODE SHARING

Upon publication, all analysis code (Python notebooks and scripts) will be uploaded to a GitHub repository.

## CLINICAL TRIAL

Not a clinical trial.

## Supplementary Materials

**Supplementary Table 1.** Videos depicting medical consultations used in this work.

| Code | Title of the Video | Country of origin of the video | Length (min) | Link to the Video |
| --- | --- | --- | --- | --- |
| v00001 | Entrevista de paciente con dolor torácico - Universidad de Piura | Peru | 14:58 | https://www.youtube.com/watch?v=evw4Ovi5v34 |
| v00002 | Modelo de Entrevista Médica - Universidad de Piura | Peru | 19:02 | https://www.youtube.com/watch?v=Y0ZSI7Pv0jQ&t=4s |
| v00003* | Vídeo de demostración de entrevista médica | Colombia | 20:19 | https://www.youtube.com/watch?v=BMAH_eBWW_w |
| v00004 | Guardia C. ( Consulta Medica ) | Mexico | 11:00 | https://www.youtube.com/watch?v=bTICaB49MwU |
| v00005 | Modelo funcional de consulta médica | Ecuador | 11:32 | https://www.youtube.com/watch?v=qM1WidnhHss |
| v00006 | Simulación de consulta médica - Shanti Granja | Ecuador | 9:40 | https://www.youtube.com/watch?v=UTO2U2xONyo |
| v00007 | SIMULACIÓN ENTREVISTA MÉDICO PACIENTE | Peru | 9:17 | https://www.youtube.com/watch?v=vfpaiJe4BGk |
| v00008 | HISTORIA CLÍNICA COMPLETA |  | 17:19 | https://www.youtube.com/watch?v=KUSecjK80wl |
| v00009 | Medical Spanish Mock Interview | USA | 13:47 | https://www.youtube.com/watch?v=b2qZOqUGe48 |
| v00010 | Entrevista Médica: Relación Médico-Paciente // UNLaM | Argentina | 6:00 | https://www.youtube.com/watch?v=ESsmoYsvtSk |

The audio recordings from the initial four videos (v00001 to v00004) were downloaded on October 20<sup>th</sup>, 2025. The audio recordings for the remaining videos were downloaded on January 26<sup>th</sup>, 2026. We provide the links where the videos were found, although readers are reminded that depending on when they read this table, some of these links may no longer be functional. The country of origin of the video was determined by the authors based on information available in the video (e.g., university name) or accent (e.g., typical Spanish accent from Argentina). This limitation should be taken into consideration. Additional meta-data from these videos were not available. We did not contact the authors or have access to any additional information beyond what is available on YouTube. Videos were retrieved through a combination of free terms in Spanish such as “*consulta medica*”, “*entrevista medica*” and “*consulta con medico*” and scrolling through the first 3-5 pages.
\*Audio left out for external validation.

**Supplementary Table 2.**
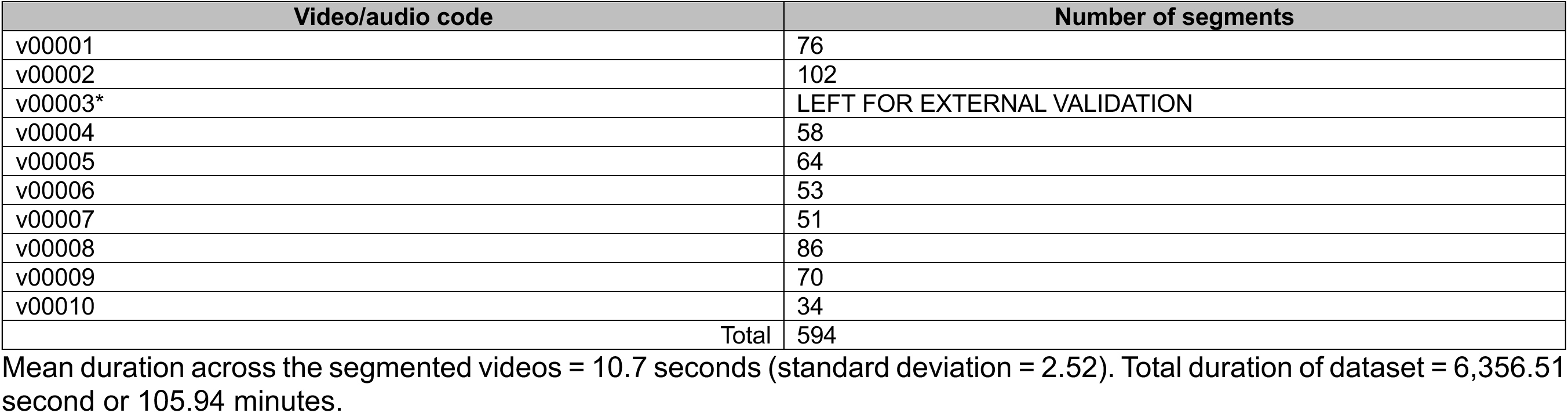
Number of segments derived from each full-length video.

| Video/audio code | Number of segments |
| --- | --- |
| v00001 | 76 |
| v00002 | 102 |
| v00003* | LEFT FOR EXTERNAL VALIDATION |
| v00004 | 58 |
| v00005 | 64 |
| v00006 | 53 |
| v00007 | 51 |
| v00008 | 86 |
| v00009 | 70 |
| v00010 | 34 |
| Total | 594 |

Mean duration across the segmented videos = 10.7 seconds (standard deviation = 2.52). Total duration of dataset = 6,356.51 second or 105.94 minutes.

## Notes

### Summary of Updates

This updated version contains all the revisions made after the first round of peer-review in a scientific journal.

